# A data-driven approach to identify clusters of HbA1c longitudinal trajectories and associated outcomes in type 2 diabetes mellitus: a large population-based cohort study

**DOI:** 10.1101/2022.06.14.22276398

**Authors:** Adrian Martinez-De la Torre, Maria Luisa Faquetti, Fernando Perez-Cruz, Christian Meier, Stefan Weiler, Andrea M. Burden

## Abstract

**Background:** We aimed to identify and characterize common patterns of HbA1c progression among type 2 diabetes mellitus patients who initiate a non-insulin antidiabetic drug (NIAD).

**Methods:** The IQVIA Medical Research Data incorporating data from THIN, a Cegedim database of anonymized electronic health records, was used to identify a cohort of patients with a first-ever prescription for a NIAD between 2006 and 2019. Trajectory clusters were identified using an Expectation-Maximization algorithm by iteratively fitting *k* thin-plate splines and reassigning each patient to the nearest cluster. Cox proportional hazards models calculated the hazard ratios (HR) and 95% confidence intervals (CI) for the estimated risk of microvascular (e.g., retinopathy, diabetic polyneuropathy [DPN]) and macrovascular events.

**Findings:** Among 116,251 new users of NIADs we found five distinct clusters of HbA1c progression, which were characterized as: optimally controlled (**OC**), adequately controlled (**AC**), sub-optimally controlled (**SOC**), poorly controlled (**PC**), and uncontrolled (**UC**). The UC and AC clusters had similar index HbA1C (>9%) but the AC cluster achieved HbA1c control (HbA1C <7.5%), while the UC cluster HbA1c remained >9.0%. Compared to the OC cluster, there was a 21% (HR: 1.21, 95% CI: 1.14-1.28) and 30% (HR: 1.30, 95% CI: 1.21-1.40) elevated risk of retinopathy in the AC and UC clusters, respectively. While the PC and UC clusters had a significant 23% (HR 1.23, 95% CI 1.12 – 1.35) and 45% (HR 1.45, 95% CI: 1.27 – 1.64) increased risk of DPN, respectively.

**Interpretation:** The five identified HbA1c trajectory clusters had different risk profiles. Despite achieving diabetic control, patients categorized in the AC cluster had similar outcomes to the UC cluster, suggesting baseline HbA1c is an important indicator of health outcomes.

**Funding:** The Swiss Data Science Centre

## Introduction

Type 2 diabetes mellitus (T2DM) is a chronic disease arising from the body’s inefficient use of insulin or progressive inability to secrete insulin, which results in abnormal blood levels of glucose.[1–3] T2DM is characterized by hyperglycemia, ultimately leading to microvascular (e.g., retinopathy) and macrovascular complications (e.g., cardiovascular disease). Disease management includes lifestyle modification and glucose control using pharmacotherapy (e.g., non-insulin antidiabetic drugs [NIADs]) to reduce diabetic complications and mortality risk.[4]

Glycated hemoglobin (HbA1c) provides a long-term trend of glucose levels in the blood over the last two to three months, and it is often used as a clinical target for glycemic control. Guidelines for disease management frequently consider HbA1c <7% (53mmol/mol) a general target for glucose control.[5–7] However, T2DM has a high degree of heterogeneity in individual patient characteristics leading to different treatment strategies and distinct treatment responses. Therefore, guidelines for T2DM management, such as the National Institute for Health and Care Excellence (NICE) in the UK, recommend a more individualized treatment approach considering patient’s preferences and individual characteristics (e.g., age, comorbidities, and multiple medications).[5]

Previous studies have shown that hyperglycemia, and therefore elevated HbA1c levels, is associated with the risk of diabetes complications and mortality.[8,9] The United Kingdom Prospective Diabetes Study (UKPDS) identified that intensive glycemic control had important long-term clinical implications on microvascular and macrovascular outcomes.[10] Using data from the Action to Control Cardiovascular Risk in Diabetes (ACCORD) trial, Wang et al. found worse cardiovascular outcomes among patients with persistent poor glycemic control, when compared to patients with HbA1c around 7%. [11] Similarly, using observational data, Liateerapong and colleagues found that 10-year glycemic control was associated with improved microvascular outcomes. [12]

Thus, identifying groups of patients with specific HbA1c courses may help to develop more personalized strategies for T2DM management. However, the evaluation of HbA1c progression with time is challenging, particularly due to patients with an unequal length of observations, unevenly spaced in time, and heterogeneous observation windows. Large data and advanced statistical models are required to identify if patient trajectories can be grouped into similar clusters, and if these trajectories affect microvascular and macrovascular outcomes. Thus, in this study we applied an Expectation-Maximization (EM) algorithm using *k*-means clustering and thin-plate splines to identify distinct HbA1c trajectories in new users of NIADs using a large population-based electronic health record database. Additionally, we evaluate the association between the trajectory clusters and subsequent microvascular and macrovascular events.

## Methods

### Data source

We used the IQVIA Medical Research Data incorporating data from THIN, a Cegedim database of anonymized electronic health records from general practitioners (GPs). The database is comprised of over 18 million patients, from 800 general practices in the UK and about 6% of the population. THIN provides detailed longitudinal information regarding patient characteristics (e.g., sex, practice registration date, and ethnicity), medical conditions (e.g., diagnoses with dates, referrals to hospitals, and symptoms), medications (e.g., generic drug name, dose, and prescription date), and additional health data (e.g., laboratory results including HbA1c, creatinine and calcium blood levels, smoking status, height, weight, alcohol use, birth and death dates).

The database contains information on drug prescriptions recorded by GPs. Medications are recorded in the database using the British National Formulary (BNF) classification, and then were mapped according to the international anatomical therapeutic codes (ATC) classification system. All diagnoses are recorded using READ codes [13], a comprehensive coding system with over 100,000 codes and comparable to the international classification of diseases (ICD) system. The study protocol was approved by the THIN scientific research council (study reference number: 20SR062).

### Study population

We identified patients aged 18+ years between January 1^st^ 2006 and December 31^st^ 2019, with new onset T2DM defined as a first-time NIAD prescription. The index date was defined as the date of the first ever NIAD prescription after start of valid data collection. All patients were required to have more than 1-year of eligible data collection. Patients with a diagnosis of polycystic ovarian syndrome or gestational diabetes prior to index date were excluded, since these conditions are often treated with NIADs. Additionally, we excluded patients with insulin prescription prior to, or at, index date and patients with less than four records of HbA1c after index date. For each the time-to-event analysis, we further excluded patients with a diagnosis of the outcome of interest previous to index.

### Study variables

We assessed variables of interest i.e., age, body mass index (BMI), smoking status, and alcohol consumption at index date. Additionally, we included the following comorbidities, defined by the presence of corresponding diagnostic or test Read codes: angina pectoris, anxiety and other neurotic, stress related and somatoform disorders, arthropathy, atrial fibrillation, cancer, chronic depression, chronic liver disease, congestive heart failure, high blood pressure, hypercholesterolemia, hypothyroidism, irritable bowel syndrome, ischemic heart disease, neuropathy, osteoarthritis, primary open-angle glaucoma, and senile cataract.

Moreover, we analyzed main laboratory results which are highly affected by T2DM, stratifying by cluster i.e., estimated glomerular filtration rate (eGFR), bilirubin, vitamin B12, serum iron, low-density lipoprotein (LDL), and triglycerides.

### Follow-up period

For each patient, there were two exposure periods. The first one was defined as starting at the date of a first-ever NIAD prescription and ending at the end of follow up (December 31^st^, 2019), death, or loss-to-follow up due to disenrollment from GP, whichever occurred first, and it was used classify the type of HbA1c trajectory.

The second exposure time started at the date of a first-ever NIAD prescription until the occurrence of any outcome of interest, ad it was used in the time-to-event analysis and the Cox-proportional hazards model.

### Outcomes of interest

The primary outcomes of interest were classified as microvascular conditions (retinopathy, diabetic polyneuropathy [DPN], and erectile dysfunction [ED]), and macrovascular diseases (acute myocardial infarction [AMI], coronary heart disease [CHD], peripheral arterial disease [PAD]). All chronic disease conditions were identified based on read codes, while insulin use was identified based on ATC codes. All included codes can be found in https://github.com/adrianmartinez-ETH/hba1c_progression.

### Statistical analysis

Analysis of longitudinal trajectories of HbA1c was conducted employing an Expectation-Maximization (EM) algorithm using *k*-means clustering and thin-plate splines (TPSs).[14] TPS are functions defined piece-wise by polynomials which are used to model relationships between a predictor *X* and a variable *Y*. The functions are fitted using a generalized additive model (GAM), as shown in **Equation 1**

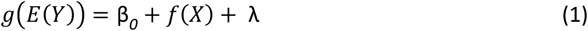

where β_0_ is a constant, *f*(*X*) a flexible function of *X*, and λ is the penalty term which constrains the function to a certain degree of smoothness. A TPS depends on the *m* data points with known coordinates and target values, and it can be described by 2(*m* + 3) parameters. Therefore, the objective is to minimize the Residual Sum of Squares (RSS) where *J* is the penalty function and λ controls the importance of this, as shown in Equation 2.

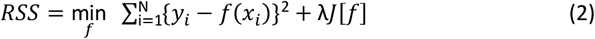

In the extreme scenario of *λ* = 0 we would fit a spline that perfectly overfits the data points, and with *λ* → ∞ we would fit the polynomial base model fitted by ordinary least squares.

TPSs provide a very flexible framework for model fitting, since it does not require any prior knowledge about the functional form, and there is no need to specify the number of nodes and their location, which allows for optimal control of continuous confounders.

We implemented an EM algorithm which allows to estimate the latent structure of the data by assuming that the observed data comes from a finite set of mixtures. We first fitted *k* different random splines and assigned each patient to the nearest cluster based on the smaller mean squared error (MSE). Then, we iteratively re-computed the *k* splines based on the clusters formed (M-step) and re-assigned group membership (E-step) until convergence. In case clusters become too small (i.e., with more observations than degrees of freedom) they may merge, resulting in a small number of clusters.

In order to select a robust number of clusters, we used three different approaches. The first approach consisted in computing a large model of 40 different clusters, followed by a hierarchical clustering analysis. The latter was performed using complete linkage based on the Euclidean distance between the fitted values of each cluster in order to visualize a potentially optimal number of clusters.[15] The second approach consisted in performing a silhouette analysis, which measured the separation between clusters, and therefore allowed for different number of potential clusters.[16] The elbow method was used in the third approach. This method was performed by computing the deviance of models of different numbers of clusters in order to visually inspect the computed the deviance of models of different sizes. The location of a bend in the plot was considered an indicator of the appropriate number of clusters.

For each cluster, we summarized the patient characteristics at index date. The history of comorbidities was identified if a valid read code was present any time prior to index. We compared patients’ characteristics at index date using t-test and chi-square tests for continuous and categorical variables, respectively, and computed the standardized mean differences (SMDs) between groups to assess the magnitude of the difference. We defined a SMD > 0.2 to indicate significance.

Time-to-event analysis was conducted for microvascular conditions (retinopathy, DPN, and ED), and for macrovascular diseases (AMI, CHD, and PAD], as well as the time to first insulin use. Kaplan-Meyer curves stratified by cluster were plotted for each outcome. Patients with a diagnosis of outcomes under investigation previous to index date were excluded from the time- to event analysis. Additionally, we fitted a Cox-proportional hazards model for each outcome adjusted for sex, age, and BMI, smoking, and alcohol consumption, at index date. Moreover, we computed the Tukey’s range test to determine if there were statistically significant differences between clusters.[17]

Finally, we explored annual changes in NIAD utilization within the first 5-years after index date. At each annual time point, we identified the proportion of patients receiving different classes of NIADs within the prior 3-month window to identify if relevant differences in NIAD utilization were present between the clusters.

## Results

### Study Population

We identified 116,251 new users of NIADs who had at least four HbA1c measurements after index date (**Figure 1**). In order to inspect for potential selection bias, we compared patient characteristics of included and excluded patients in this study (**Supplementary Table S1)**. No substantial differences between included and excluded patients were observed.

**Figure 1.**
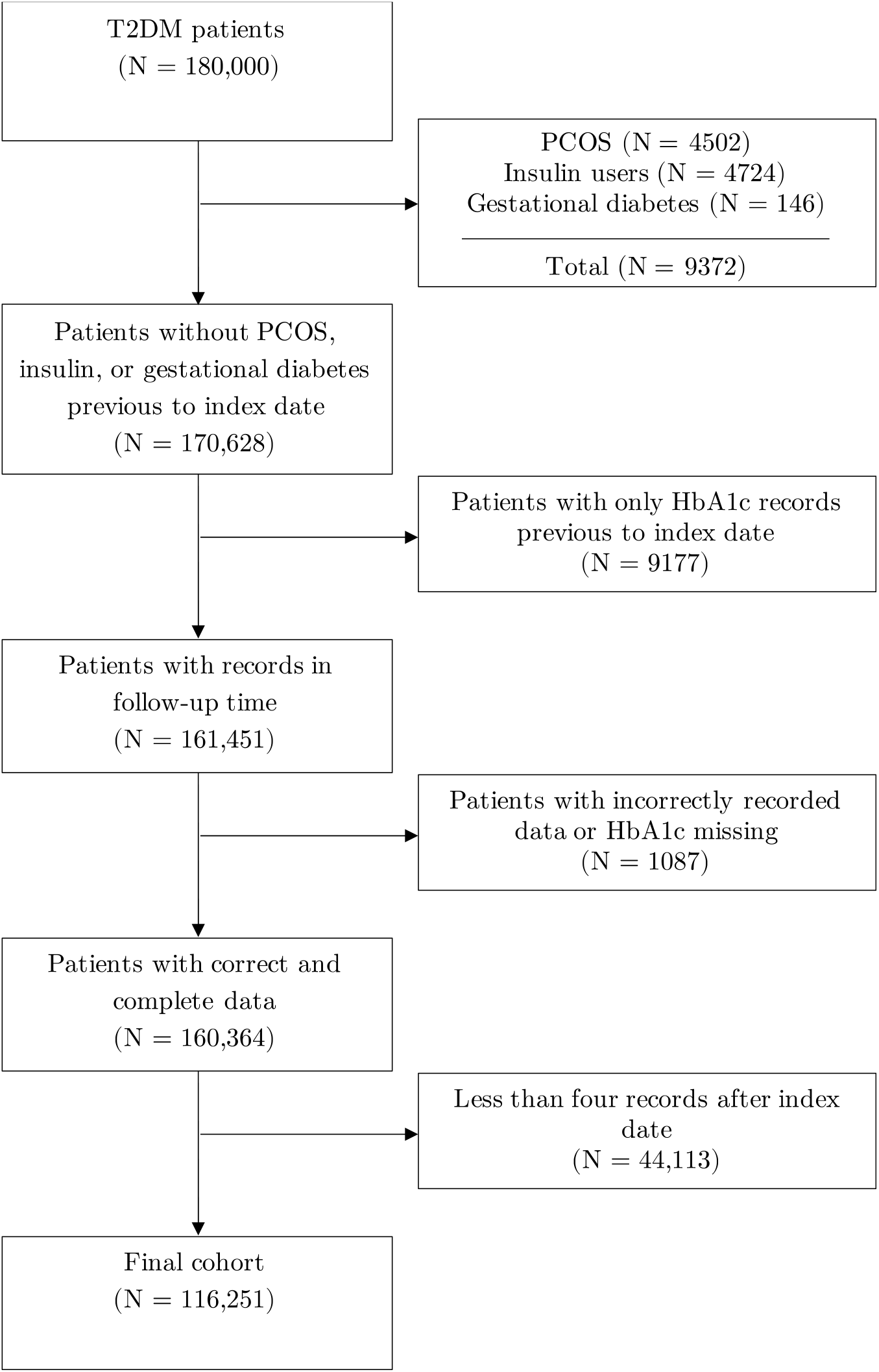
Flowchart of included patients.

### Longitudinal HbA1c trajectories

The three different approaches (hierarchical clustering analysis, the silhouette analysis, and the elbow method) used to select a robust number of clusters identified an optimal number of clusters between four and six clusters. Given that four clusters were too general and did not manage to capture and represent all the patterns, and six clusters provided little benefit at the expense of overfitting, we opted for five clusters, **Supplementary Figure S1**.

**Figure 2** provides a visual depiction of the cluster trajectories. While all clusters showed an initial drop in HbA1c levels following NIAD start, HbA1c trajectories varied greatly between clusters. When characterizing the five clusters we identified the following patterns:

1. Optimally controlled HbA1c (**OC**),
2. Adequately controlled HbA1C (**AC**),
3. Suboptimally controlled HbA1c (**SOC**)
4. Poorly controlled HbA1c (**PC**)
5. Uncontrolled HbA1c (**UC**)

**Figure 2.**
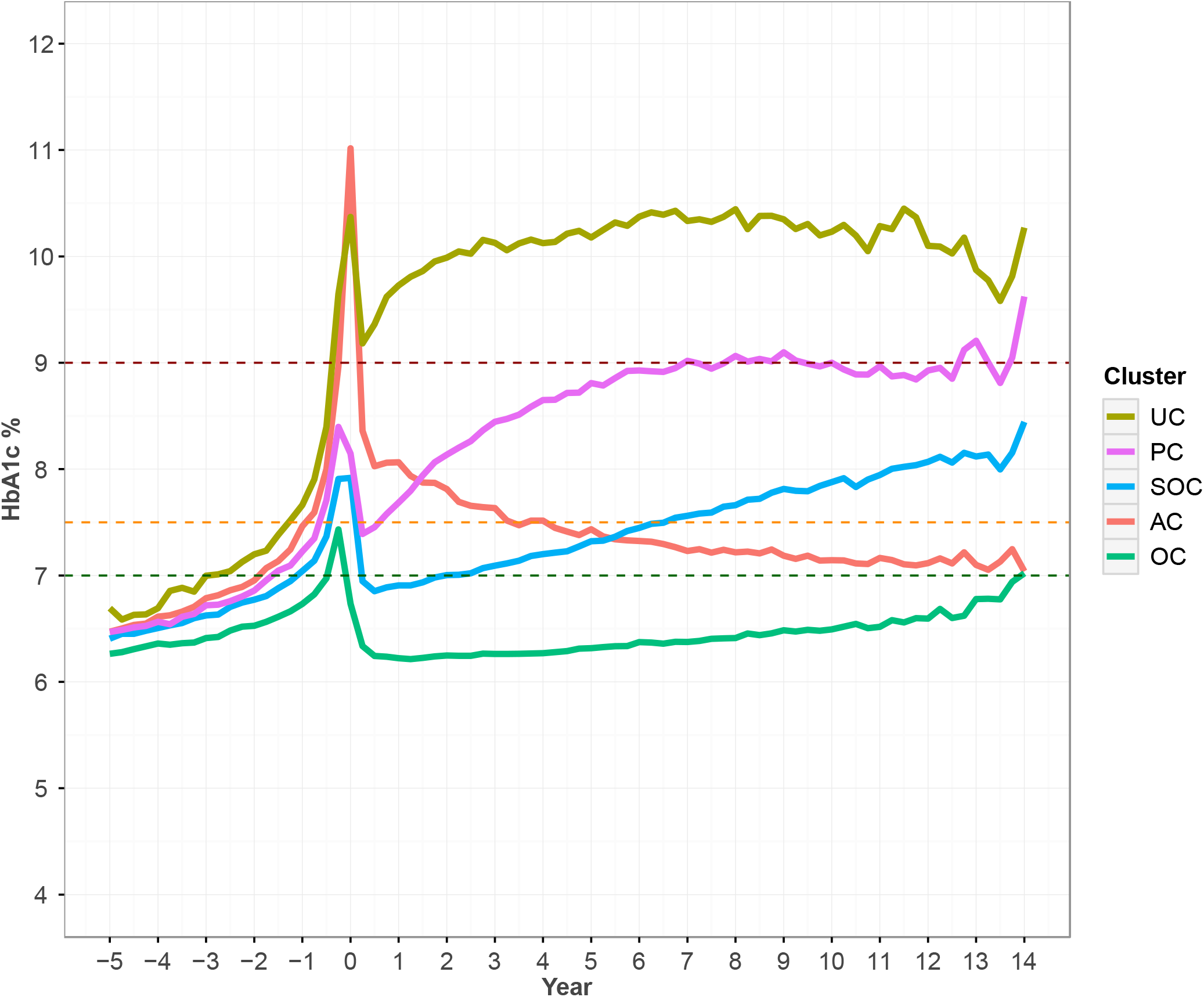
Evolution of cluster centroids of HbA1c level in percentage. **Abbreviations:** UC, uncontrolled HbA1; PC, poor HbA1c control; SOC, suboptimal HbA1c control; AC, adequate HbA1c response; OC, optimal HbA1c control. The gray area indicates the start of NIAD therapy (index date), the green, orange, and red horizontal dashed lines indicate an optimal control (≤7%), adequate HbA1c level after treatment intensification (≤ 7.5%), and level where insulin should be considered (≥ 9%), respectively.

The first OC cluster (28.8%; n=33,531) had a modest decrease in HbA1c from baseline and HbA1c levels that remained mostly below 7.0%. The second AC cluster (14.6%; n=16,962) showed a significant initial reduction in HbA1c after index and HbA1c levels remained below the clinical target of 7.5% from year 4 onward. Although patients in the third SOC cluster (32.1%; n=37,325) had an initial drop in HbA1c levels a gradual increased over time was noted, and by year 10 was above 7.5% and on par with the baseline HbA1c. The fourth PC cluster (17.1%; n=19,832) had a moderate initial HbA1c reduction, followed by a rapid increase, with the 10-year HbA1c reaching 9.0% and above the baseline value. And finally, patients in the fifth UC cluster (7.4%; n=8,601) never achieved HbA1c control, with values remaining above 9.0% and reaching above 10% by year 10.

Patient characteristics at index date, stratified by cluster, are shown in **Table 1**. The AC and UC clusters started with the highest HbA1c values (>9%), yet the long-term trajectories were strikingly different, thus, we provide the significance test between the AC and UC clusters in Table 1. Patients in the UC cluster were significantly younger when compared to the AC cluster (53.8 years vs. 60.5 years, SMD 0.53), had a higher mean BMI (34.1 vs. 32.6, SMD 0.21), and were more likely to be current smokers (26.6% vs. 18.2%, SMD 0.22). The UC group had a lower prevalence of high blood pressure (34.3% vs. 42.9%, SMD 0.18) compared to the AC cluster. Conversely, the UC cluster had the highest proportion of patients with chronic liver disease (3.1%) and anxiety and stress (21.3%). Nonetheless, although there were statistically significant differences between both groups, the SMDs were not significant for comorbidities.

**Table 1.** Patient characteristics at index date, stratified by cluster.

|  | **OC**<br>(N=33531) | **AC**<br>(N=16962) | **SOC**<br>(N=37325) | **PC**<br>(N=19832) | **UC**<br>(N=8601) | **AC vs UC**<br>**P value** | **AC vs UC**<br>**SMD** |
| --- | --- | --- | --- | --- | --- | --- | --- |
| Gender = Male (%) | 18091 (54.0) | 10376 (61.2) | 21463 (57.5) | 11756 (59.3) | 5091 (59.2) | 0.002 | 0.04 |
| Age (mean (standard deviation)) | 63.8 (12.4) | 60.5 (12.0) | 62.7 (12.0) | 58.2 (12.8) | 53.8 (13.0) | <0.001 | 0.53 |
| BMI (mean (standard deviation)) | 32.5 (6.8) | 32.6 (6.9) | 32.2 (6.4) | 33.4 (6.9) | 34.1 (7.6) | <0.001 | 0.21 |
| Alcohol - Current use (%) | 23061 (72.9) | 11502 (73.7) | 25742 (73.4) | 13467 (72.9) | 5465 (70.0) | <0.001 | 0.08 |
| Smoking - Current use (%) | 4845 (14.5) | 3076 (18.2) | 5597 (15.0) | 3904 (19.8) | 2269 (26.6) | <0.001 | 0.22 |
| Conditions |  |  |  |  |  |  |  |
| Angina pectoris | 2802 (8.8) | 1048 (6.8) | 2920 (8.3) | 1188 (6.4) | 339 (4.4) | <0.001 | 0.10 |
| Anxiety & other* | 5828 (18.4) | 2596 (16.7) | 6126 (17.4) | 3574 (19.3) | 1644 (21.3) | <0.001 | 0.12 |
| Arthropathy | 2385 (7.5) | 903 (5.8) | 2338 (6.6) | 1094 (5.9) | 365 (4.7) | <0.001 | 0.05 |
| Atrial fibrillation | 2685 (8.5) | 1183 (7.6) | 2715 (7.7) | 1374 (7.4) | 468 (6.1) | <0.001 | 0.06 |
| Cancer | 9330 (29.5) | 3977 (25.6) | 9968 (28.3) | 4731 (25.6) | 1730 (22.4) | <0.001 | 0.02 |
| Chronic depression | 199 (0.6) | 92 (0.6) | 210 (0.6) | 132 (0.7) | 67 (0.9) | <0.001 | 0.03 |
| Chronic liver disease | 852 (2.7) | 407 (2.6) | 915 (2.6) | 551 (3.0) | 243 (3.1) | <0.001 | 0.03 |
| Congestive heart failure | 1016 (3.2) | 524 (3.4) | 1076 (3.1) | 543 (2.9) | 228 (2.9) | <0.001 | 0.02 |
| High blood pressure | 15889 (50.2) | 6651 (42.9) | 16414 (46.6) | 7662 (41.4) | 2652 (34.3) | <0.001 | 0.18 |
| Hypercholesterolaemia | 6831 (21.6) | 2726 (17.6) | 7187 (20.4) | 3125 (16.9) | 1061 (13.7) | <0.001 | 0.11 |
| Hypothyroidism | 3215 (10.2) | 1303 (8.4) | 3163 (9.0) | 1556 (8.4) | 670 (8.7) | <0.001 | 0.01 |
| Intermittent claudication | 1262 (4.0) | 526 (3.4) | 1293 (3.7) | 581 (3.1) | 184 (2.4) | <0.001 | 0.06 |
| Irritable bowel syndrome | 3541 (11.2) | 1403 (9.0) | 3735 (10.6) | 1951 (10.5) | 713 (9.2) | <0.001 | 0.01 |
| Ischaemic heart disease | 560 (1.8) | 244 (1.6) | 634 (1.8) | 310 (1.7) | 105 (1.4) | <0.001 | 0.02 |
| Neuropathy | 321 (1.0) | 115 (0.7) | 303 (0.9) | 135 (0.7) | 47 (0.6) | <0.001 | 0.02 |
| Osteoarthritis | 8035 (25.4) | 3247 (20.9) | 8287 (23.5) | 3646 (19.7) | 1204 (15.6) | <0.001 | 0.00 |
| Primary open-angle glaucoma | 1479 (4.7) | 513 (3.3) | 1422 (4.0) | 568 (3.1) | 165 (2.1) | <0.001 | 0.07 |
| Senile cataract | 1998 (6.3) | 696 (4.5) | 1888 (5.4) | 786 (4.2) | 217 (2.8) | <0.001 | 0.03 |

**Abbreviations:** UC, uncontrolled HbA1; PC, poor HbA1c control; SOC, suboptimal HbA1c control; AC, adequate HbA1c response; OC, optimal HbA1c control; BMI, body mass index; Anxiety & other\*, anxiety and other neurotic, stress related, and somatoform disorders.

**Table 2.**
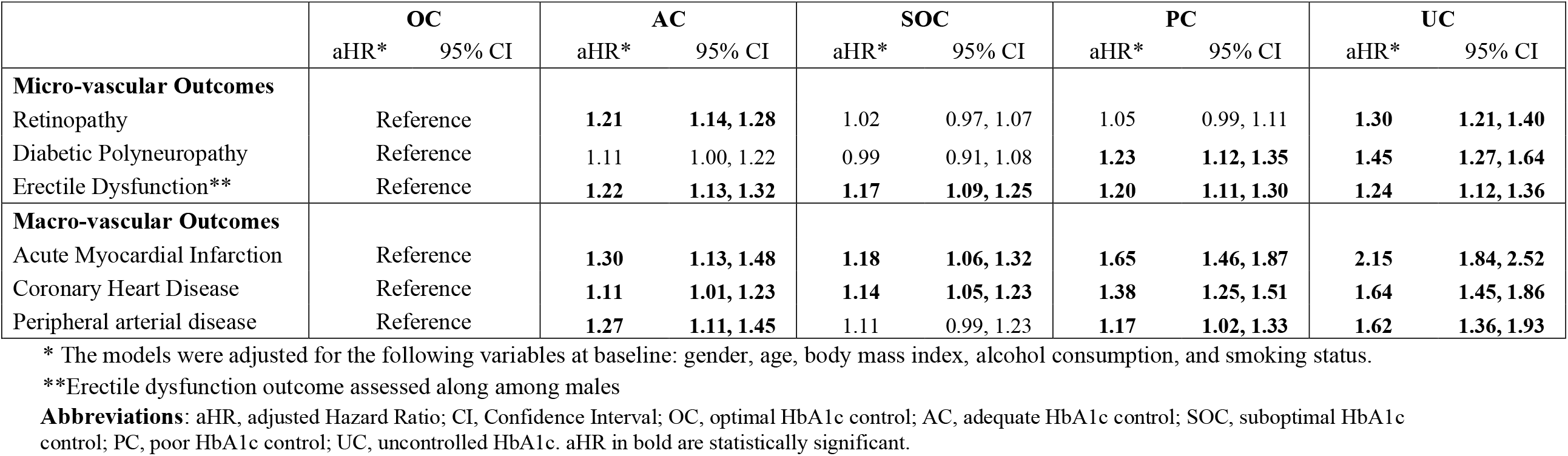
Adjusted hazard ratio of the different outcomes.

Among the other clusters, we note that patients in the OC cluster were more frequently older (average age of 63.8 years old) and less likely to be smokers (14.5%) when compared to other clusters, **Table 1**. On the other hand, this group had the highest proportion of patients with a history of high blood pressure (50.2%), hypercholesterolemia (21.6%), and osteoarthritis (25.4%). The SOC and PC clusters both showed increasing HbA1c trajectories over time, and were relatively similar at baseline. The SOC cluster was slightly older (62.7 years vs. 58.2 years) than the PC cluster, and slightly less likely to be current smokers (15.0% vs. 19.8%). The SOC cluster had the lowest average BMI (32.2 kg/m^2^) compared to other groups.

We observed that patients in the UC and PC clusters had overall good kidney (normal values: eGFR ≥ 60 mL/min/1.73 m^2^) and liver function (normal values: bilirubin ≤ 21 μmol/L), as well as good vitamin B12 levels (normal values: ≥ 200 and ≤ 950 pg/mL). On the other hand, these patients presented with the highest levels of LDL (normal values: ≥ 2.6 and ≤ 4.1) and triglycerides (normal values: ≥ 1.69 and ≤ 2.25 mmol/mol), **Supplementary Figure S2**.

### Risk for microvascular and macrovascular outcomes

The adjusted Cox-proportional hazards models for the microvascular and macrovascular outcomes of interest are provided in **Table 3**, and the unadjusted survival curves and number of events are provided in **Supplementary Figure S3** and **Supplementary Table S2**, respectively. For retinopathy, only the AC and UC clusters presented statistically significant hazard ratios (HRs) with a 21% (HR 1.21, 95% CI: 1.14 – 1.28) and 30% (HR 1.30, 95% CI: 1.21 – 1.40) increased risk with respect to the OC cluster, respectively. When assessing DPN, the PC and UC clusters had a significant 23% (HR 1.23, 95% CI 1.12 – 1.35) and 45% (HR 1.45, 95% CI: 1.27 – 1.64) increased risk, respectively, when compared to the OC group. All clusters showed an approximate 20% significant increased risk of ED, when compared to the OC cluster. Between cluster differences are presented in **Supplementary Table S3**. There were no statistically significant differences between the AC and UC clusters for retinopathy risk (p=0.312), nor between the PC and UC clusters for DPN risk (p=0.100). Similarly, no between group differences were noted for ED.

For macrovascular events, all clusters showed an increased risk of AMI, compared to the OC group, with the highest risk observed among the UC cluster (HR 2.15, 95% CI: 1.84–2.52), **Table 3**. When comparing the AC vs. the UC cluster with the Tukey’s range test we found statistically significant differences between each other (p<0.001), **Supplementary Table S3**. For CHD, all the clusters showed an elevated risk when compared to the OC cluster, with the highest risk again being among the UC cluster (HR 1.64, 95% CI: 1.45–1.86). While for PAD, the AC and UC clusters showed a 27% (HR 1.27, 95% CI: 1.11–1.45) and 62% (HR 1.62, 95% CI: 1.36–1.93) increased risk, respectively, compared to the OC cluster, but no statistically significant differences between each other.

### Prescription patterns

Over 90% of the patients started treatment at index date with biguanides e.g., metformin, followed by sulfonylureas, **Supplementary Figure S4**. Nonetheless, patients in the UC cluster were the first ones to stop biguanides treatment the earliest and have them replaced the earliest by other medications. This cluster was the one with the highest proportion of patients with insulins, SGLT2 inhibitors, and GLP-1 at the five-years mark. The AC cluster, who had a similar initial trajectory but a dramatic improvement with respect to the UC cluster, was the group with the greatest number of users of thiazolidinediones (TZDs).

## Discussion

In this population-based cohort study, we identified five distinct patterns of HbA1c progression with different patient and clinical risk profiles. We found two clusters (UC and AC) with similar high HbA1c values at the start of NIAD therapy, but with very different trajectories. Nevertheless, similar microvascular and macrovascular risks were observed among both groups, when compared to the OC cluster suggesting high baseline HbA1c may be an important risk factor. Additionally, we found that the clusters with the highest risk of AMI and CHD were observed among the clusters with elevated HbA1c during follow-up, the PC and UC clusters.

In addition to this, the UC cluster presented the highest use of insulin in the first five years after NIAD start. Further work should aim to assess if patient-level predictors of cluster assignment can be identified to aide in optimizing treatment and management strategies.

In the last twenty years there has been an increase in the prevalence of T2DM worldwide, linked to a more sedentary lifestyle, diet, and an increasing aging population.[18] Moreover, T2DM and its complications have contributed substantially to the global burden of mortality and disability, as diabetes is one of the major causes of reduced life expectancy.[19] Thus, improving our understanding the evolution patterns of HbA1c levels at T2DM diagnosis is paramount to advancing tailored therapeutic management in order to achieve glycemic control.

While most of the studies that have aimed at modelling HbA1c progression have used latent class growth modelling (LCGM) with smaller cohorts and shorter follow-up periods,[11,12,20] we identified similar trajectory patterns. For example, we found that deteriorating HbA1c (i.e.,, PC) and extremely high baseline HbA1c (i.e., AC and UC) were associated with worse clinical outcomes, when compared to the cluster with stable values (i.e., OC).

To date, only the work of Laiteerapong et al. used a LCGM in combination of a larger real-world cohort of 28,016 individuals.[21] This study identified five distinct HbA1c trajectories, and found an association between non-stable trajectories and greater risk of microvascular events and mortality. While we looked at individual outcomes, rather than composite endpoints, our results are comparable for microvascular events. The AC cluster was similar to the “high decreasing early” cluster in the Laiteerapong study, which was associated with a 28% increased risk of microvascular events.[12] In our analyses we could see differences between the different microvascular events, which could not be observed in the Laiteerapong study. For example, while the UC cluster had elevated risks for all outcomes, the AC cluster was only significantly associated with diabetic retinopathy and ED, while the PC cluster was associated with a significant increased risk of DPN and ED. As DPN typically develops 10-20 years after the initial diabetes diagnosis, it is plausible that long-term diabetes control is an important predictor. Conversely, retinopathy typical emerges within the first 5-years, and therefore, early glycemic control is likely a key predictor.

While we did not cluster based on patient characteristics, we could identify distinct patient profiles across the five clusters. For example, the OC cluster was older and included patients with a history of age-related comorbidities at index, while patients in the worst performing group (UC) were overall younger and more frequently obese. Additionally, some differences between the AC and UC cluster were identified, namely that the AC cluster was older, had a lower mean BMI, and were less frequently smokers. As we identified differences in the likelihood of clinical outcomes, particularly for the AC and UC clusters, and two previous studies found that data-driven clusters based on baseline characteristics (including HbA1c) may be predictive of clinical outcomes.[22,23] Future work should further investigate the patient-level factors that are predictive of the cluster orientation.

Finally, many guidelines for diabetes management are moving towards individualized treatment to improve long-term glucose control and clinical outcomes. We could identify some differences between the clusters regarding the treatment course. For example, the UC cluster moved to second-line therapies earliest, especially insulins. Interestingly, the AC cluster appeared to have the highest proportion of users switching to TZDs early in therapy. These results are preliminary, but provide insight into the potential for tailored therapy in T2DM.

When interpreting our results, we have to acknowledge several limitations. First of all, we only looked at patients after the start of their first NIAD prescription, thus we might have missed patterns of patients that were first instructed to change their diet and physical activity levels in order to attain glycemic control. Additionally, we were restricted to data from the UK where intrinsic social factors such as lifestyle, diet, or physical exercise might have a relevant impact in glycemic control. In addition, treatment guidelines have changed several times during our follow-up period i.e., 2009 and 2015. Therefore, the relevant thresholds considered to attain a controlled glycemic status have been revised and updated, as well as the medications used. Although metformin still remains the first-line therapy, several types of drugs have been developed, approved, and marketed during our study window. For instance, SGLT2 inhibitors such as dapaglifozin or canaglifozin were approved in 2012 and 2013 respectively. The fact that we excluded patients with less than four HbA1c measurements after index date might could have introduced a selection bias in favor of either healthier or more careless patients. Nonetheless, we did not find relevant differences between included and excluded patients overall, as shown in **Supplementary Table S1**. A potential limitation in the methodological approach we used of splines in combination with *k*-means clustering is the fact that we had to empirically select the number of trajectories. Although we minimized the potential source of bias by performing three different methods i.e., elbow method, hierarchical clustering, and silhouette analysis, we did not find clear cutoff points for the number of clusters. Finally, since we included patients with a first NIAD prescription between January 2003 to December 2019, not many patients have a follow-up period of more than ten years. This led to a higher variance and volatility in the last years of the time-to-event analyses.

In conclusion, we found that clusters with worse baseline and long-term glycemic control were associated with higher degree long term microvascular and macrovascular complications. Moreover, as we identified that high baseline HbA1c, as seen in the UC and AC clusters, maybe a strong indicator of retinopathy risk, while long-term HbA1c control is associated with DPN. Further studies should aim at understanding the differences between clusters of similar profile but diverging trajectories, and investigate if more tailored therapy can help improve long-term glycemic trajectories in patients with T2DM.

## Data Availability

The IQVIA Medical Research Data (IMRD) were obtained from IQVIA, a Cegedim database of anonymized electronic health records. For further information on access to the database, please contact IQVIA (contact details can be found at https://www.iqvia.com/locations/united-kingdom/information-for-members-of-the-public/medical-research-data).

## Author Contributions

Study Conception: AMB, FPC; data acquisition: AMB; data analysis: AMDlT; data integrity and validity: AMDlT, AMB; data interpretation: AMDlT, FPC, MLF, CM, SW, AMB; manuscript preparation: AMDlT, MLF, AMB; critical revisions: AMDlT, MLF, CM, FPC, SW, AMB.

## Ethics statement

The protocol for this project was approved by the THIN scientific research council (reference number: 20SR062).

## Consent to participate and for publication

Consent to participate and for publication was granted by the THIN scientific research council (reference number: 20SR062).

