## Supplementary Material for "A data-driven approach to identify clusters of HbA1c longitudinal trajectories and associated outcomes in type 2 diabetes mellitus: a large population-based cohort study"

**Title**

### Author Names and Affiliations

Adrian Martinez-De la Torre[1], M.Sc

Maria Luisa Faquetti[1], M.Sc

Fernando Perez-Cruz[2,3], Ph.D

Christian Meier[4], MD

Stefan Weiler[1,5], MD, Ph.D

Andrea M. Burden[1], Ph.D

[1] Institute of Pharmaceutical Sciences, Department of Chemistry and Applied Biosciences, ETH Zurich, Zurich, Switzerland.
[2] Swiss Data Science Center, ETH Zurich and EPFL, Switzerland
[3] Institute of Machine Learning in the Computer Science Department at ETH Zurich
[4] Department of Endocrinology, Diabetology and Metabolism, University Hospital Basel, Basel, Switzerland.
[5] Clinical Pharmacology and Toxicology, Department of General Internal Medicine, Inselspital, Bern University Hospital, University of Bern, Bern, Switzerland

### Corresponding Author

Prof. Dr. Andrea Burden

Institute of Pharmaceutical Sciences, Department of Chemistry and Applied Biosciences

ETH Zurich

Vladimir-Prelog-Weg 1-5/10

8093 Zurich

Switzerland

**Supplementary Table S1.** Patient characteristics at index date between included and excluded patients.

| | **INCLUDED** (N=116251) | **EXCLUDED** (N=44113) | **p-value** | **SMD** |
|---|---|---|---|---|
| Gender = Male (%) | 66777 (57.4) | 23927 (54.2) | <0.001 | 0.06 |
| Age (mean (standard deviation)) | 61.26 (12.66) | 60.58 (15.48) | <0.001 | 0.05 |
| BMI (mean (standard deviation)) | 32.67 (6.77) | 32.62 (7.27) | 0.266 | 0.01 |
| Alcohol – Current (%) | 79237 (73.0) | 28497 (69.5) | <0.001 | 0.08 |
| Smoking – Current (%) | 19691 (17.0) | 7999 (18.2) | <0.001 | 0.05 |
| Conditions | | | | |
| Angina pectoris | 8297 (7.6) | 2732 (6.8) | <0.001 | 0.03 |
| Anxiety & other* | 19768 (18.2) | 8158 (20.3) | <0.001 | 0.05 |
| Arthropathy | 7085 (6.5) | 2530 (6.3) | 0.129 | 0.01 |
| Atrial fibrillation | 8425 (7.8) | 3774 (9.4) | <0.001 | 0.06 |
| Cancer | 29736 (27.4) | 12485 (31.1) | <0.001 | 0.08 |
| Chronic Depression | 700 (0.6) | 274 (0.7) | 0.439 | 0.00 |
| Chronic liver disease | 2968 (2.7) | 1520 (3.8) | <0.001 | 0.06 |
| Congestive heart failure | 3387 (3.1) | 1690 (4.2) | <0.001 | 0.06 |
| High blood pressure | 49268 (45.4) | 16639 (41.5) | <0.001 | 0.08 |
| Hypercholesterolaemia | 20930 (19.3) | 7137 (17.8) | <0.001 | 0.04 |
| Hypothyroidism | 9907 (9.1) | 3608 (9.0) | 0.436 | 0.00 |
| Intermittent claudication | 3846 (3.5) | 1532 (3.8) | 0.012 | 0.01 |
| Irritable bowel syndrome | 11343 (10.4) | 4682 (11.7) | <0.001 | 0.04 |
| Ischaemic heart disease | 1853 (1.7) | 647 (1.6) | 0.218 | 0.01 |
| Neuropathy | 921 (0.8) | 403 (1.0) | 0.005 | 0.02 |
| Osteoarthritis | 24419 (22.5) | 8905 (22.2) | 0.225 | 0.01 |
| Primary open-angle glaucoma | 4147 (3.8) | 1697 (4.2) | <0.001 | 0.02 |
| Senile cataract | 5585 (5.1) | 2493 (6.2) | <0.001 | 0.05 |

**Abbreviations**: SMD, standardized mean difference; anxiety and other, anxiety and other neurotic, stress related and somatoform disorder; BMI, body mass index.

**Supplementary Table S2.** Average time to event of selected outcomes.

| | **OC** (N=33,531) | **AC** (N=16,962) | **SOC** (N=37,325) | **PC** (N=19,832) | **UC** (N=8601) | **p-value** |
|---|---|---|---|---|---|---|
| **Retinopathy** | | | | | | |
| N | 3541 (10.6) | 2168 (12.8) | 4071 (10.9) | 2160 (10.9) | 1114 (13.0) | |
| Time to event (mean (SD)) | 35.3 (31.5) | 39.3 (34.8) | 37.3 (32.9) | 41.5 (36.7) | 46.5 (39.7) | <0.001 |
| **Diabetic peripheral neuropathy** | | | | | | |
| N | 1185 (3.5) | 648 (3.8) | 1286 (3.4) | 782 (3.9) | 345 (4.0) | |
| Time to event (mean (SD)) | 36.41 (31.1) | 37.85 (31.0) | 39.31 (32.9) | 44.54 (36.9) | 45.00 (35.8) | <0.001 |
| **Erectile dysfunction** | | | | | | |
| N | 1563 (8.6) | 1228 (11.8) | 2241 (10.4) | 1408 (12.2) | 701 (13.8) | |
| Time to event (mean (SD)) | 29.14 (28.0) | 28.88 (27.0) | 31.59 (29.5) | 34.41 (31.7) | 36.65 (31.6) | <0.001 |
| **AMI** | | | | | | |
| N | 634 (1.9) | 390 (2.3) | 803 (2.2) | 516 (2.6) | 259 (3.0) | |
| Time to event (mean (SD)) | 46.22 (37.2) | 48.29 (38.2) | 44.96 (35.9) | 48.65 (37.2) | 52.83 (37.1) | 0.033 |
| **Coronary heart disease** | | | | | | |
| N | 1161 (3.5) | 643 (3.8) | 1452 (3.9) | 853 (4.3) | 407 (4.7) | |
| Time to event (mean (SD)) | 42.17 (34.9) | 43.0 (35.3) | 41.7 (35.2) | 44.7 (35.9) | 47.4 (36.8) | 0.03 |
| **Peripheral arterial disease** | | | | | | |
| N | 662 (2.0) | 383 (2.3) | 780 (2.1) | 387 (2.0) | 196 (2.3) | |
| Time to event (mean (SD)) | 40.91 (33.1) | 43.89 (35.2) | 41.88 (33.6) | 50.15 (38.4) | 50.72 (38.6) | <0.001 |

**Abbreviations:** N, number of cases; OC, optimal HbA1c control; AC, adequate HbA1c control; SOC, suboptimal HbA1c control; PC, poor HbA1c control; UC, uncontrolled HbA1c.

**Supplementary Table S3.** Contrasts of clusters from Cox Proportional Hazard models between clusters.

| Microvascular Outcomes | | | | | | | | | | | | | |
|---|---|---|---|---|---|---|---|---|---|---|---|---|---|
| | | Retinopathy | | | | Diabetic Neuropathy | | | | Erectile Dysfunction | | | |
| Comparison groups | | Estimate | Std. Error | z-value | p-value | Estimate | Std. Error | z-value | p-value | Estimate | Std. Error | z-value | p-value |
| AC | OC | 0.19 | 0.03 | 6.54 | <0.001 | 0.10 | 0.05 | 1.95 | 0.284 | 0.20 | 0.04 | 4.97 | <0.001 |
| SOC | OC | 0.02 | 0.02 | 0.87 | 0.905 | -0.01 | 0.04 | -0.22 | 0.999 | 0.15 | 0.03 | 4.47 | <0.002 |
| PC | OC | 0.04 | 0.03 | 1.55 | 0.521 | 0.20 | 0.05 | 4.23 | <0.001 | 0.18 | 0.04 | 4.73 | <0.003 |
| UC | OC | 0.26 | 0.04 | 7.03 | <0.001 | 0.37 | 0.07 | 5.69 | <0.001 | 0.21 | 0.05 | 4.28 | 0.000 |
| SOC | AC | -0.17 | 0.03 | -5.96 | <0.001 | -0.11 | 0.05 | -2.16 | 0.188 | -0.05 | 0.04 | -1.26 | 0.709 |
| PC | AC | -0.14 | 0.03 | -4.46 | <0.001 | 0.10 | 0.06 | 1.87 | 0.328 | -0.02 | 0.04 | -0.40 | 0.995 |
| UC | AC | 0.08 | 0.04 | 1.90 | 0.312 | 0.27 | 0.07 | 3.83 | 0.001 | 0.01 | 0.05 | 0.23 | 0.999 |
| PC | SOC | 0.02 | 0.03 | 0.85 | 0.911 | 0.21 | 0.05 | 4.51 | <0.001 | 0.03 | 0.04 | 0.86 | 0.908 |
| UC | SOC | 0.24 | 0.04 | 6.60 | <0.001 | 0.38 | 0.06 | 5.91 | <0.001 | 0.06 | 0.05 | 1.25 | 0.716 |
| UC | PC | 0.22 | 0.04 | 5.54 | <0.001 | 0.16 | 0.07 | 2.44 | 0.100 | 0.03 | 0.05 | 0.57 | 0.979 |
| Macrovascular Outcomes | | | | | | | | | | | | | |
| | | Acute Myocardial Infarction | | | | Coronary Heart Disease | | | | Peripheral Arterial Disease | | | |
| Comparison groups | | Estimate | Std. Error | z-value | p-value | Estimate | Std. Error | z-value | p-value | Estimate | Std. Error | z-value | p-value |
| AC | OC | 0.26 | 0.07 | 3.81 | 0.001 | 0.11 | 0.05 | 2.10 | 0.216 | 0.24 | 0.07 | 3.54 | 0.004 |
| SOC | OC | 0.17 | 0.06 | 3.05 | 0.019 | 0.13 | 0.04 | 3.15 | 0.014 | 0.10 | 0.06 | 1.85 | 0.337 |
| PC | OC | 0.50 | 0.06 | 8.02 | <0.001 | 0.32 | 0.05 | 6.72 | <0.001 | 0.15 | 0.07 | 2.26 | 0.154 |
| UC | OC | 0.77 | 0.08 | 9.49 | <0.001 | 0.50 | 0.06 | 7.94 | <0.001 | 0.48 | 0.09 | 5.47 | <0.001 |
| SOC | AC | -0.09 | 0.07 | -1.39 | 0.630 | 0.02 | 0.05 | 0.42 | 0.993 | -0.14 | 0.07 | -2.10 | 0.215 |
| PC | AC | 0.24 | 0.07 | 3.45 | 0.005 | 0.21 | 0.05 | 3.86 | <0.001 | -0.09 | 0.08 | -1.12 | 0.790 |
| UC | AC | 0.51 | 0.09 | 5.86 | <0.001 | 0.39 | 0.07 | 5.73 | <0.001 | 0.24 | 0.09 | 2.60 | 0.067 |
| PC | SOC | 0.33 | 0.06 | 5.64 | <0.002 | 0.19 | 0.05 | 4.21 | <0.001 | 0.05 | 0.07 | 0.79 | 0.932 |
| UC | SOC | 0.60 | 0.08 | 7.67 | <0.003 | 0.37 | 0.06 | 6.06 | <0.001 | 0.38 | 0.09 | 4.41 | <0.001 |
| UC | PC | 0.26 | 0.08 | 3.24 | 0.010 | 0.18 | 0.06 | 2.76 | 0.044 | 0.33 | 0.09 | 3.52 | 0.004 |

**Supplementary Figure S1.** Silhouette analysis from $k$=3 to $k$=8, elbow method from $k$=1 to $k$=10, and hierarchical clustering of 40 clusters with $k$=5, Euclidian distance, and complete linkage.

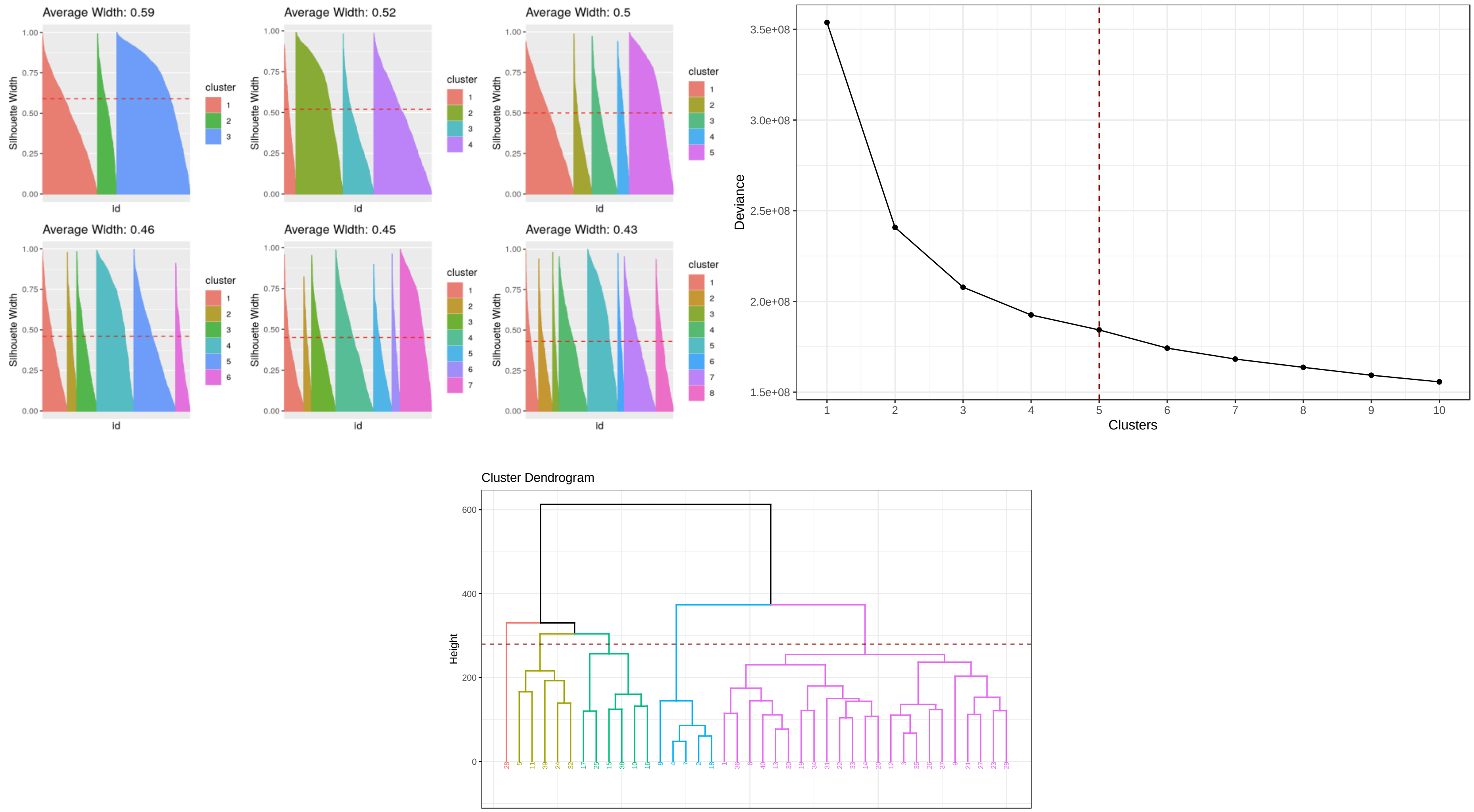

**Supplementary Figure S2.** Evolution of main lab results by cluster.

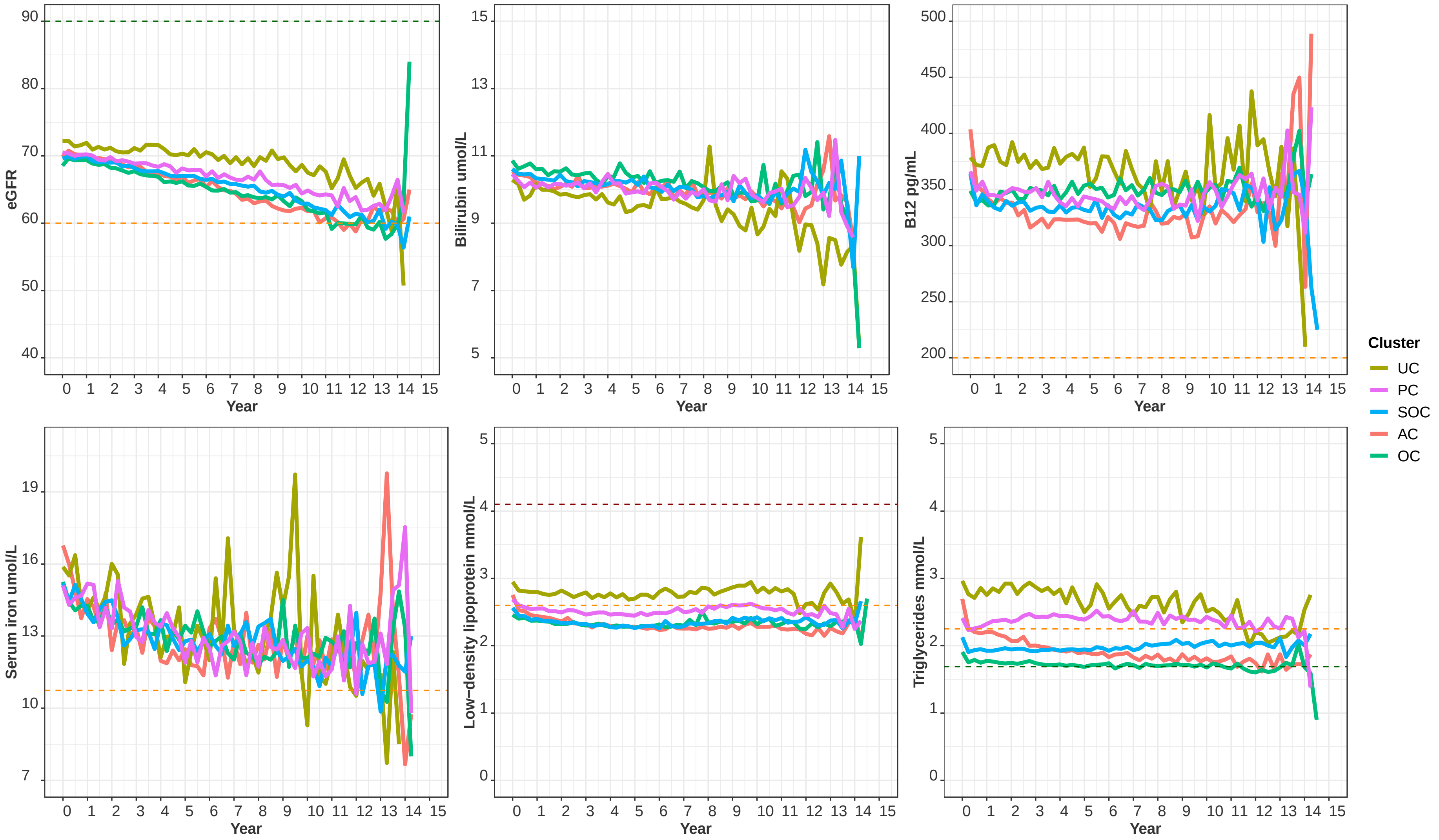


**Abbreviations:** eGFR, estimated glomerular filtration rate; OC, optimal HbA1c control; AC, adequate HbA1c control; SOC, suboptimal HbA1c control; PC, poor HbA1c control; UC, uncontrolled HbA1c.

**Supplementary Figure S3.** Cumulative number of cases for each of the selected comorbidities stratified by cluster.

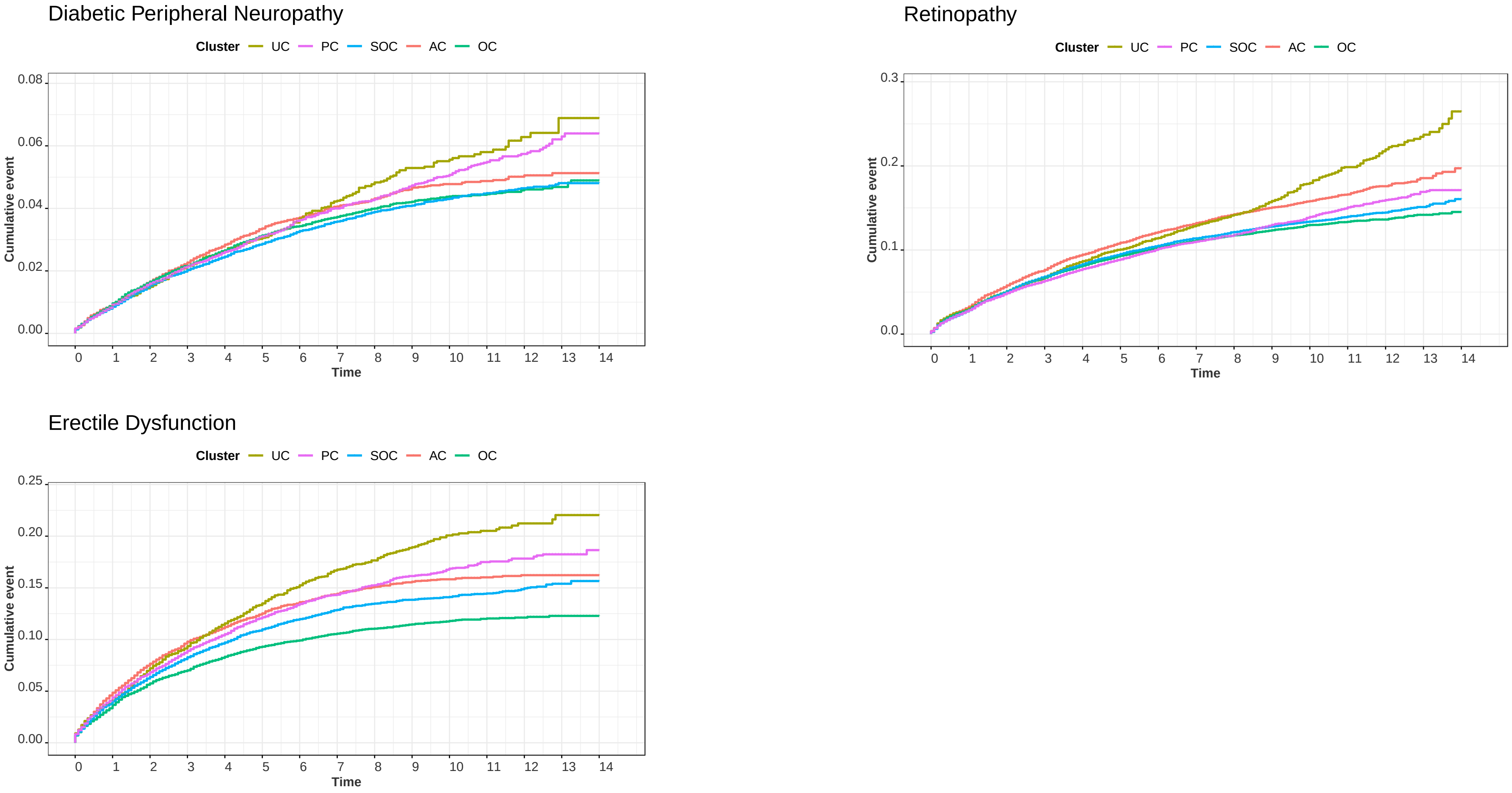


**Abbreviations:** OC, optimal HbA1c control; AC, adequate HbA1c control; SOC, suboptimal HbA1c control; PC, poor HbA1c control; UC, uncontrolled HbA1c.

**Supplementary Figure S3. (Continued)** Cumulative proportion of cases for each of the selected comorbidities stratified by cluster.

Coronary Heart Disease

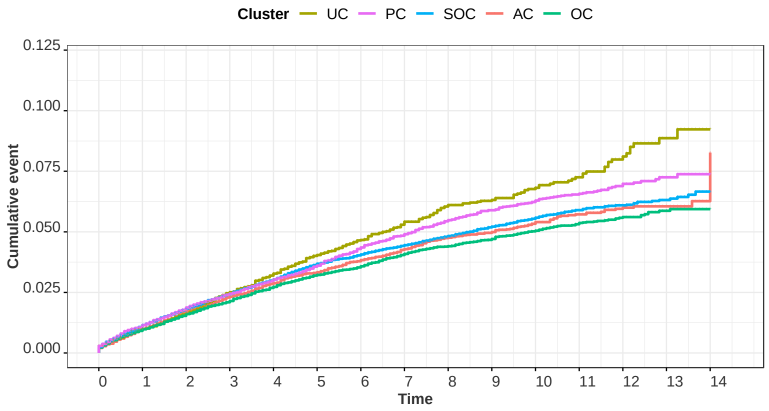


Acute Myocardial Infarction

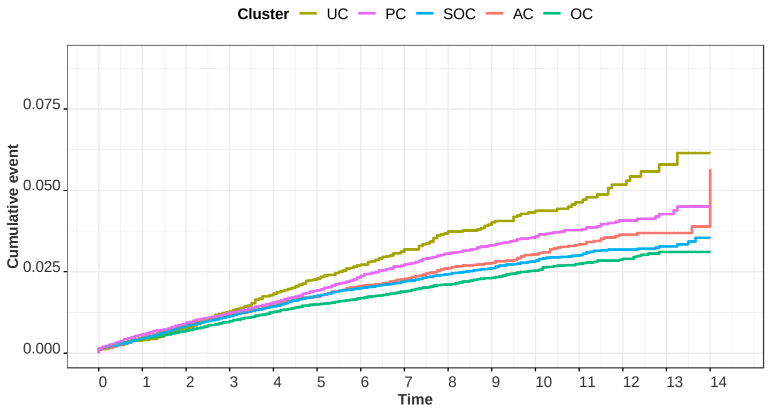


Peripherial Arterial Disease

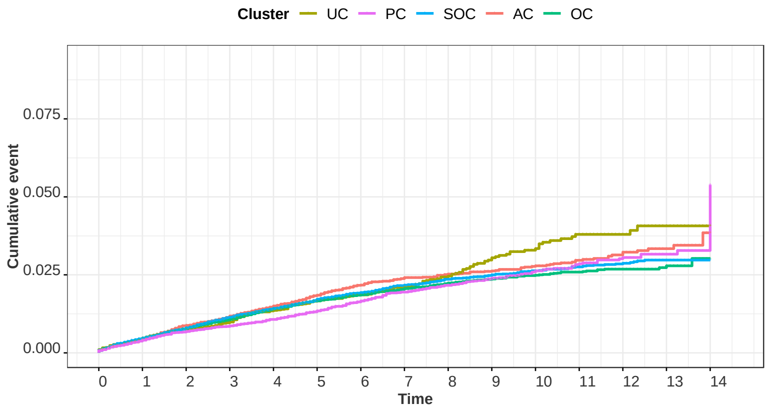


**Abbreviations:** OC, optimal HbA1c control; AC, adequate HbA1c control; SOC, suboptimal HbA1c control; PC, poor HbA1c control; UC, uncontrolled HbA1c.

**Supplementary Figure S4.** Evolution of prescribed antidiabetic medications in the first five years after NIAD start.

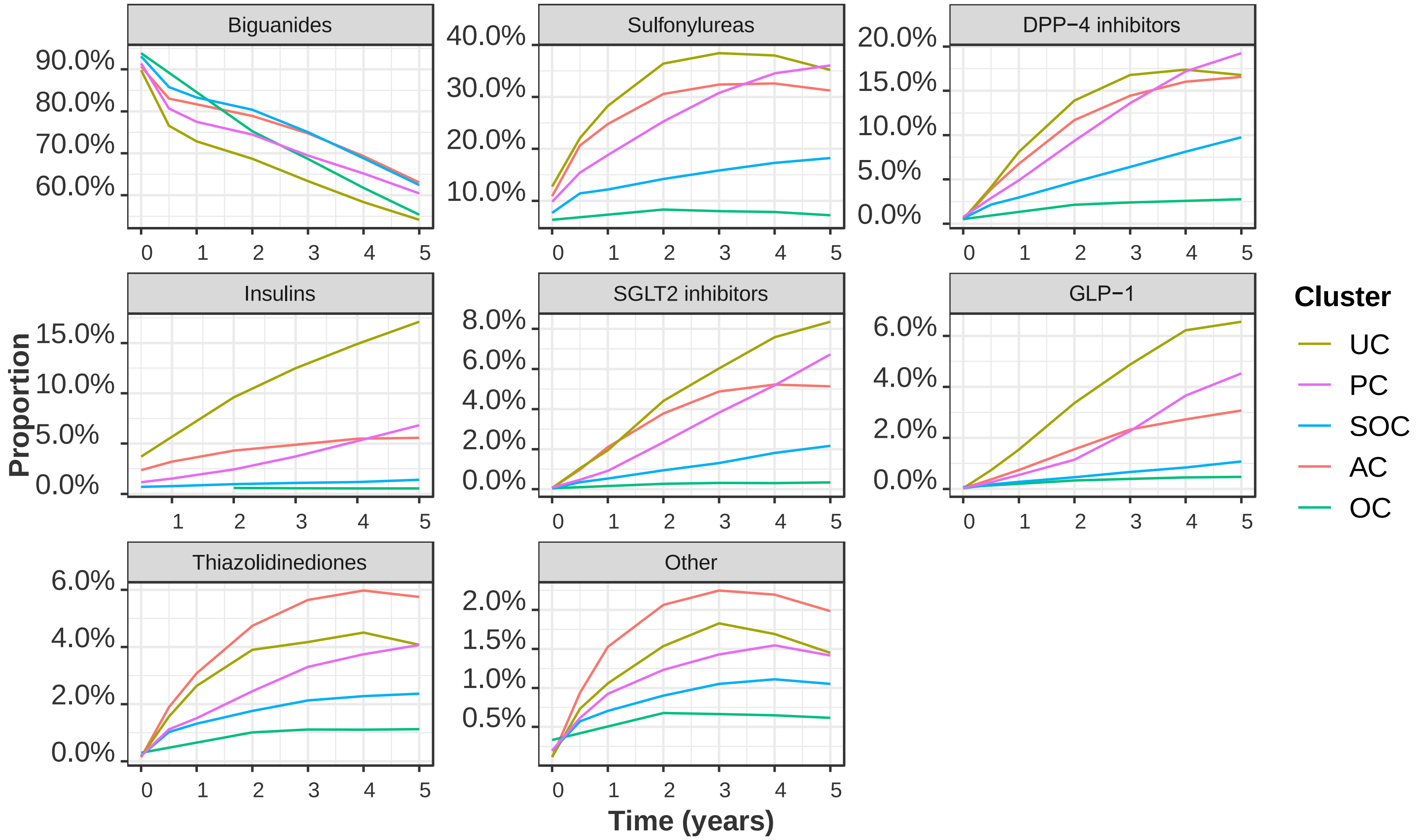


**Abbreviations:** UC, uncontrolled HbA1; PC, poor HbA1c control; SOC, suboptimal HbA1c control; AC, adequate HbA1c response; OC, optimal HbA1c control; SMD, standardized mean difference. Insulins do not include the medication at baseline since patients with insulin at index date were excluded as shown in **Figure 1**.
